# Testicular pain after living kidney donation: Results from a multicenter cohort study

**DOI:** 10.1101/2024.12.21.24319490

**Authors:** the Donor Nephrectomy Outcomes Research Network, Amit X. Garg, Liane S. Feldman, Jessica M. Sontrop, Meaghan S. Cuerden, Jennifer B. Arnold, Neil Boudville, Martin Karpinski, Scott Klarenbach, Greg Knoll, Charmaine E. Lok, Eric McArthur, Matthew Miller, Mauricio Monroy-Cuadros, Kyla L. Naylor, G V Ramesh Prasad, Leroy Storsley, Christopher Nguan

**Author notes:** **Corresponding Author**: Amit X. Garg, MD, PhD, Victoria Hospital, 800 Commissioners Rd, Room ELL-200, London, ON, Canada N6A 5W9.

## Abstract

**Background:** Some men who donate a kidney have reported testicular pain after donation; however, attribution to donation is not clear as no prior studies included a comparison group of nondonors.

**Objective:** To examine the proportion of male donors who reported testicular pain in the years after nephrectomy compared to male nondonors with similar baseline health characteristics.

**Design, Participants, and Setting:** We enrolled 1042 living kidney donors (351 male) before nephrectomy from 17 transplant centers (12 in Canada and 5 in Australia) from 2004–2014. A concurrent sample of 396 nondonors (126 male) was enrolled. Follow-up occurred until November 2021.

**Measurements:** Donors and nondonors completed the same schedule of measurements at baseline (before nephrectomy) and follow-up. During follow-up, participants completed a questionnaire asking whether they had experienced new pain in their eyes, hands, or testicles; those who experienced pain were asked to indicate on which side of the body the pain occurred (left or right). The pain questionnaire was completed by 290 of 351 male donors (83%) and 97 of 126 male nondonors (77%) a median of 3 years after baseline (interquartile range 2, 6).

**Methods:** Inverse probability of treatment weighting on a propensity score was used to balance donors and nondonors on baseline characteristics. After weighting, the nondonor sample increased to a pseudo sample of 295, and most baseline characteristics were similar between donors and nondonors.

**Results:** At baseline, donors (n=290) were a mean age of 49 years; 83% were employed, and 80% were married; 246 (84.8%) underwent laparoscopic surgery and 44 (15.2%) open surgery; 253 (87.2%) had a left-sided nephrectomy and 37 (12.8%) a right-sided nephrectomy. In the weighted analysis, the risk of testicular pain was significantly greater among donors than nondonors: 51/290 (17.6%) vs. 7/295 (2.3%); weighted risk ratio, 7.8 (95% CI, 2.7 to 22.8). Donors and nondonors did not differ statistically in terms of self-reported eye pain or hand pain. Among donors, the occurrence of testicular pain was most often unilateral (92.2%) and on the same side as the nephrectomy (90.2%). Testicular pain occurred more often in donors who had laparoscopic vs. open surgery: 48/246 (19.5%) vs. 3/44 (6.8%) but was similar in those who had a left-sided vs. right-sided nephrectomy: 44/253 (17.4%) vs. 7/37 (18.9%).

**Limitations:** Participants recalled their symptoms several years after baseline, and we did not assess the timing, severity, or duration of pain or any treatments received for the pain.

**Conclusion:** Unilateral testicular pain on the same side of a nephrectomy is a potential complication of living kidney donation that warrants further investigation.

**Clinicaltrial.gov record:** NCT00936078

## BACKGROUND

Transplantation is vital for millions of people with kidney failure as it offers a longer and better quality of life compared to dialysis and at a lower cost.^1,2^ Due to a shortage of kidneys from deceased donors, many countries strive to increase their rates of living kidney donation. Ongoing monitoring for any complications and long-term risks in living donors is essential for this practice.

To help improve our understanding of outcomes attributable to donation, we conducted a multicenter prospective cohort study of living kidney donors and nondonors of similar baseline health.^3^ We enrolled donors and nondonors from Canada and Australia between 2004 and 2014, and follow-up for study outcomes occurred until November 2021.^4–8^ While conducting this study, we became aware of reports suggesting that some men who donate a kidney experience testicular pain after donation, most often unilateral on the same side as their nephrectomy. We identified 12 studies published since 2003 that have described this outcome.^9–20^ The follow-up duration of many studies was limited, and no study included a comparison group of nondonors. In two studies, testicular pain was more common in donors who had a laparoscopic vs. open nephrectomy;^12,14^ however, most studies only examined this outcome in donors who had laparoscopic nephrectomies.^9,10,13,15–17,19,20^ The pain has been described as dull, aching, and heavy, starting approximately within one week following the nephrectomy. Only two studies compared this outcome in donors who had a left vs. right nephrectomy (with no difference observed);^12,17^ however, some have suggested this complication may be less common after a right-sided nephrectomy where, due to anatomy, the gonadal vein may be spared, whereas in a left nephrectomy, it is divided.^13,17^

After learning about this potential complication, we added a simple pain questionnaire to the follow-up survey in our cohort study, with questions about new testicular pain experienced during follow-up. Participants completed the questionnaire a median of 3 (interquartile range 2–6) years after cohort entry. Here, we describe the proportion of male donors who reported testicular pain during follow-up compared to male nondonors with similar baseline health characteristics. Among donors, we also examined whether the occurrence of testicular pain was unilateral or bilateral, whether it occurred on the same side as the nephrectomy (ipsilateral), and whether it differed between those who had a laparoscopic vs. open nephrectomy or a left-sided vs. right-sided nephrectomy.

## METHODS

### Study Design and Setting

This multicenter prospective cohort study was conducted at 17 transplant centers (12 in Canada and 5 in Australia). Participants (living kidney donors and nondonors) were recruited from 2004 to 2014, including a pilot phase from 2004 to 2008. Follow-up occurred until November 2021. The study protocol has been published (ClinicalTrials.gov NCT00936078).^3^ Ethics approval was obtained from the University of Western Ontario’s Health Sciences Research Ethics Board (REB approval 6056) and all other recruiting centers. Study conduct and reporting follow recommended STROBE guidelines (eTable 1).^21^ All participants provided written informed consent at the time of enrolment.

### Participants

Prospective donors were invited to participate in the study by their nephrologist or a coordinator at the transplant evaluation center and enrolled before surgery.^3^ A concurrent sample of nondonors was recruited from the study transplant centers. Nondonors included family members and friends of prospective donors, some individuals who came forward for donation but did not proceed with further evaluation or surgery despite being eligible, and some who came forward based on clinic advertisements. Nondonors were carefully screened to ensure their baseline health was similar to donors; nondonor eligibility criteria included age 18–70 years, body mass index (BMI) <35 kg/m^2^, systolic/diastolic blood pressure <140/90 mm Hg, and no comorbidities considered contraindications to living kidney donation (the complete list of eligibility criteria is in eTable 2).

### Data Collection and Measures

Donors and nondonors were assigned the same schedule of baseline and follow-up measurements. At baseline, all participants completed a standardized health questionnaire, had their blood pressure, height, and weight assessed, and completed lab testing for serum creatinine, urine protein, and hematuria.^3^ The donors’ nephrectomy dates and surgical details were obtained from hospital records. After they completed their baseline assessment, nondonors were assigned a simulated nephrectomy date, which served as their cohort entry date.^4^

During follow-up, participants completed a mailed survey with a simple pain questionnaire that included questions on whether the person had experienced new testicular pain after the nephrectomy (or the cohort entry date in nondonors). To assess the potential for recall bias, the questionnaire also asked about pain experienced in the hands or eyes, which we did not expect to differ between donors and nondonors. Those who experienced pain were asked to indicate on which side of the body the pain occurred (left or right) (Appendix). Participants were also asked to “please describe the pain they feel or felt.” Participants completed the questionnaire a median of 3 years after baseline (interquartile range 2, 6).

### Statistical Analysis

We used SAS software (version 9.4; SAS Institute, Inc) for the statistical analysis. Continuous variables are presented as means (standard deviations [SDs]) or medians (interquartile ranges [IQRs]), as appropriate, and categorical variables as frequencies (percentages). We used inverse probability of treatment weighting on the propensity score to balance donors and nondonors on baseline characteristics. The propensity score was estimated in a logistic regression model where the probability of being a donor vs. a nondonor was conditional on the following baseline variables: age at nephrectomy (age at cohort entry in nondonors)), self-reported history of depression diagnosed by a physician, self-reported history of anxiety diagnosed by a physician, prior kidney stones, estimated glomerular filtration rate (eGFR), and BMI. Nondonors were weighted using average treatment effect in the treated weights (where *treatment* refers to donation), defined as propensity score/(1-propensity score), with donors receiving weights of 1.

This method produced a larger weighted pseudo sample of nondonors with a similar distribution of measured baseline characteristics as donors. Between-group differences in baseline characteristics were examined using standardized differences.^22^ Many characteristics that differed between donors and nondonors (standardized mean differences > 10%) were not clinically meaningful with respect to the study outcome (they would not be expected to influence the occurrence of testicular pain). All relevant comparative analyses used methods to account for the weighting. Weighted risk differences and 95% confidence intervals (CI) were computed using binomial regression with identity link, and weighted risk ratios and 95% CIs were computed using modified Poisson regression.^23^ Bootstrapping was used to calculate confidence intervals for risk differences, and robust standard errors were used to calculate confidence intervals for risk ratios.

In addition, we conducted unweighted adjusted analyses, which controlled for the following baseline characteristics: age at nephrectomy/cohort entry, medical history of depression, medical history of anxiety, kidney stones, eGFR, and BMI. We compared these effect estimates to those obtained from the weighted analysis. Nondonors were the referent group in donor-to-nondonor comparisons. For analyses within donors, we considered the outcomes of any testicular pain and ipsilateral testicular pain.

Among donors, we compared the proportion who had testicular pain after laparoscopic vs. open surgery, and we considered whether the results were consistent after adjusting for the side of nephrectomy (left vs. right). We also examined differences in testicular pain between those who had a left-vs. right-sided nephrectomy, and we examined whether the results were consistent when stratified by laparoscopic vs. open nephrectomy. Risk differences were estimated from a binomial regression model with an identity link, and risk ratios from a modified Poisson regression model.^23^ We also examined the occurrence of reported testicular pain across surgical centers and by year of nephrectomy. Finally, we examined whether pre-donation age, height, weight, or BMI differed between donors who reported testicular pain in follow-up and those who did not; mean values were compared using a t-test.

## RESULTS

We enrolled 1042 living kidney donors before surgery (of whom 351 were male) and a concurrent sample of 396 nondonors (of whom 126 were male) (**Figure 1**). Male donors and nondonors who completed the pain questionnaire during follow-up (290 donors [83%] and 97 nondonors [77%]) were included in the analysis. At baseline, donors had a mean age of 49 years and a mean BMI of 27 kg/m^2^; 93% were white, 80% were married, 83% were employed, and 16% were smokers (**Table 1**).

**Figure 1.**
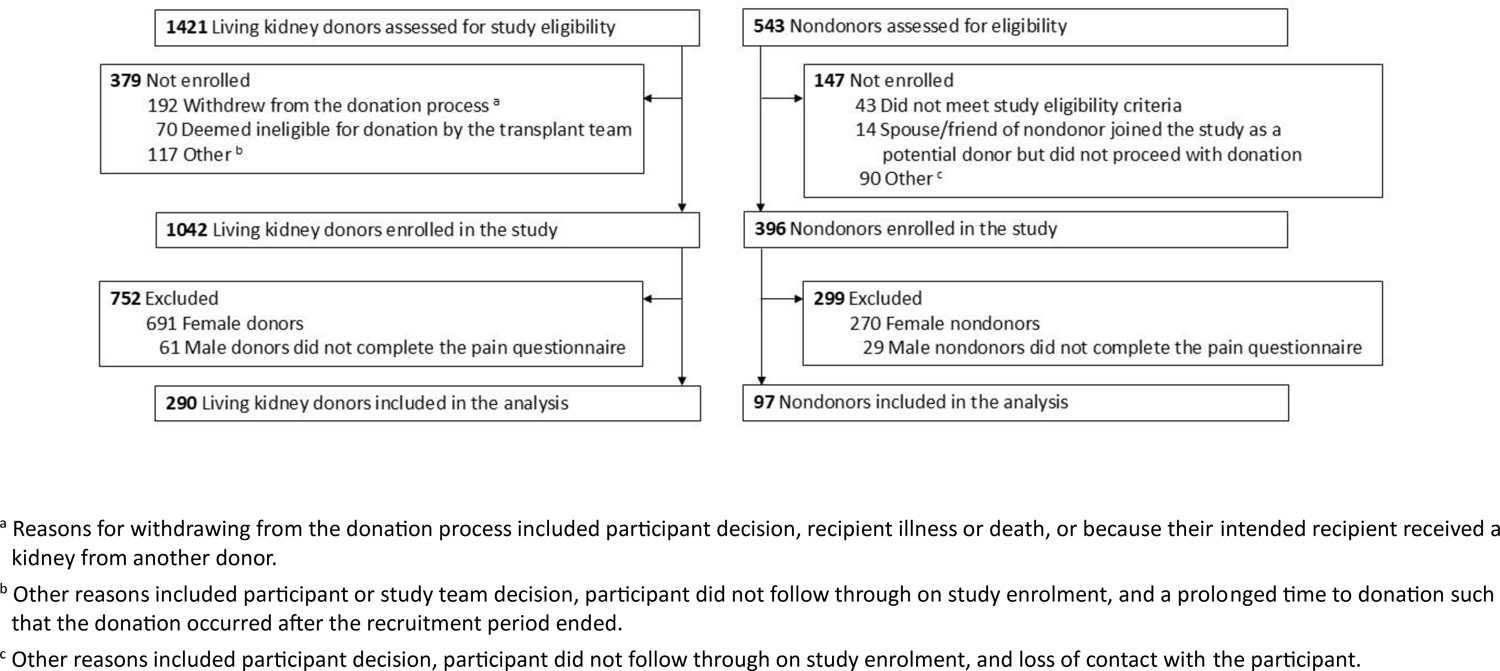
Flow diagram

**Table 1.**
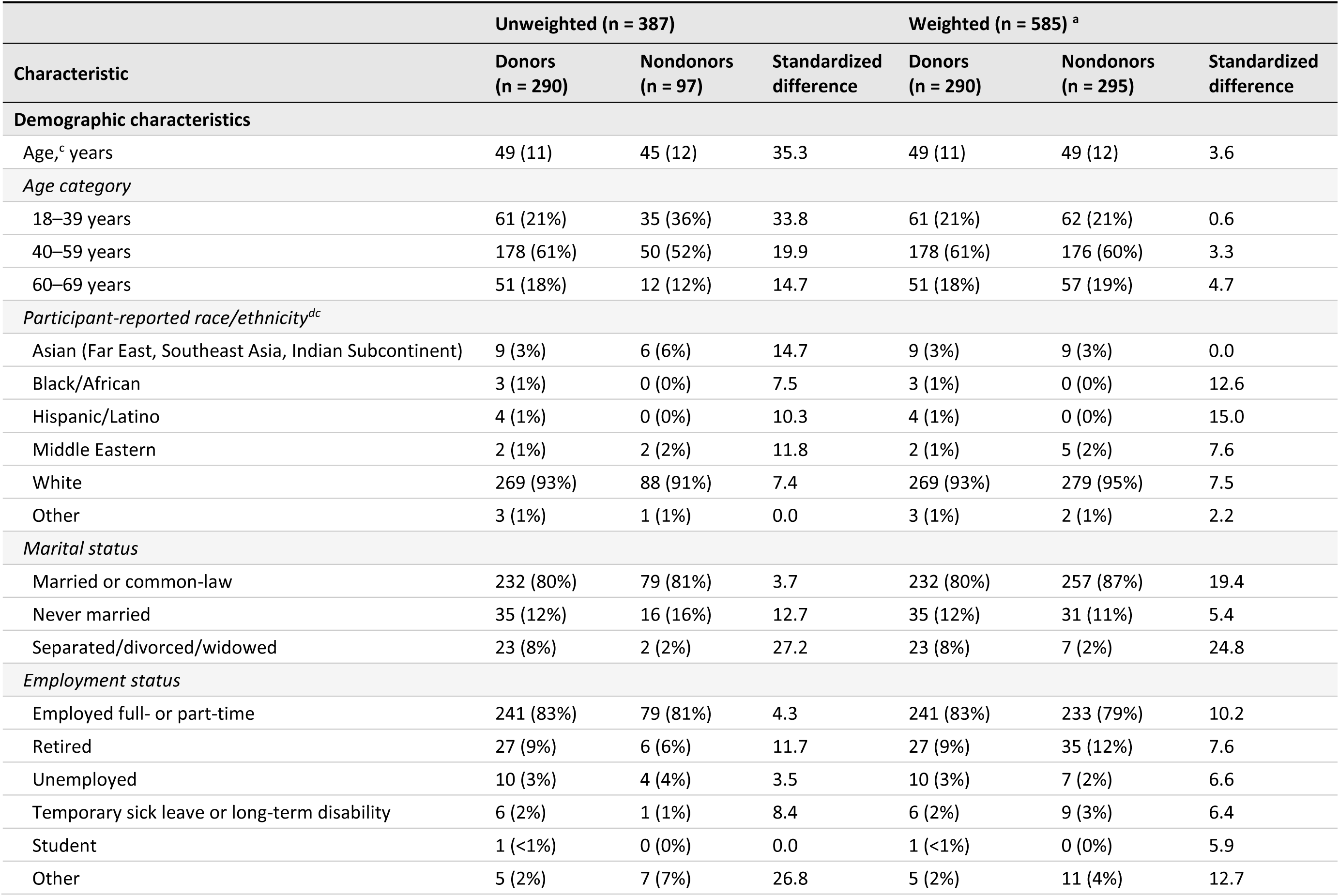
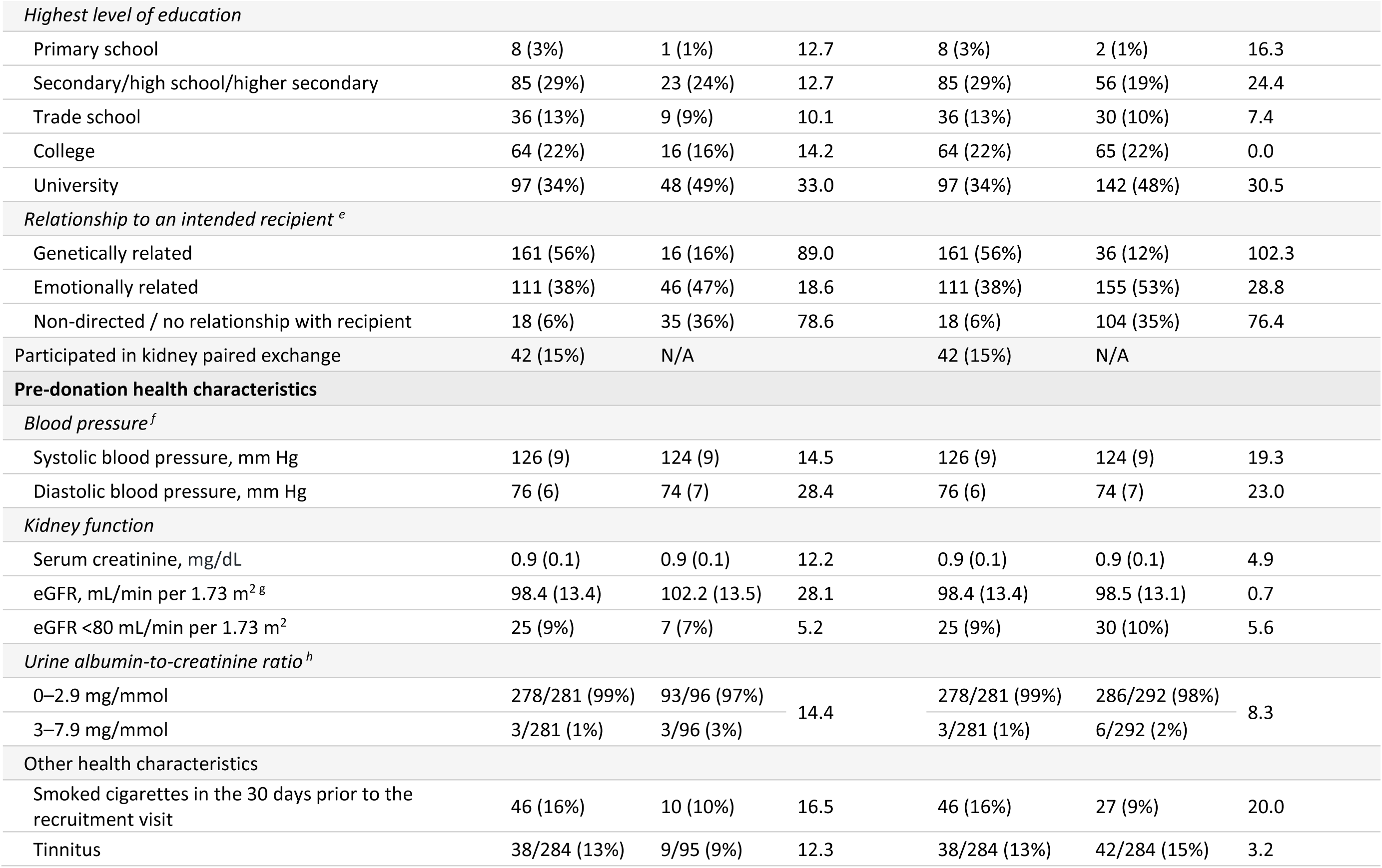
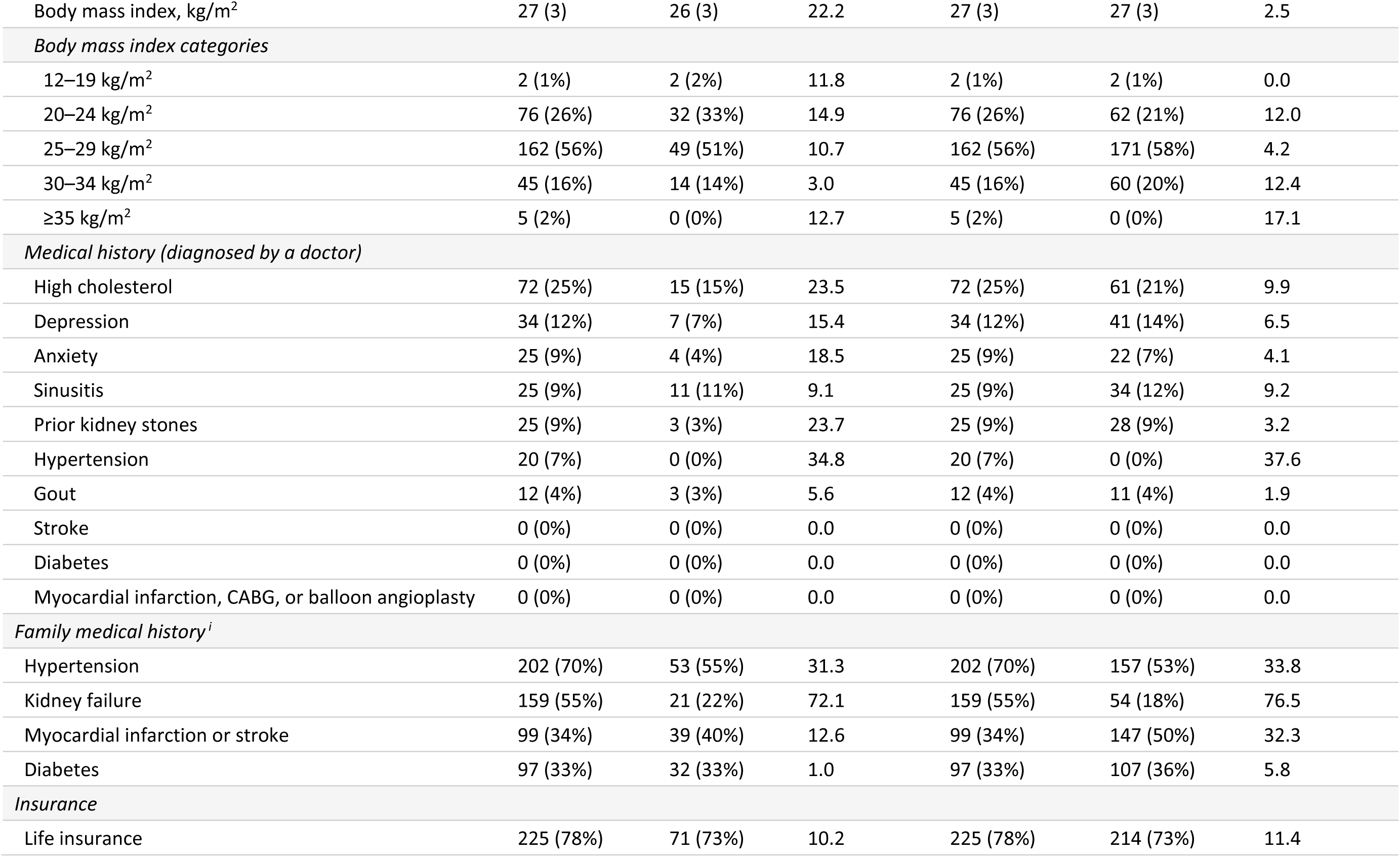
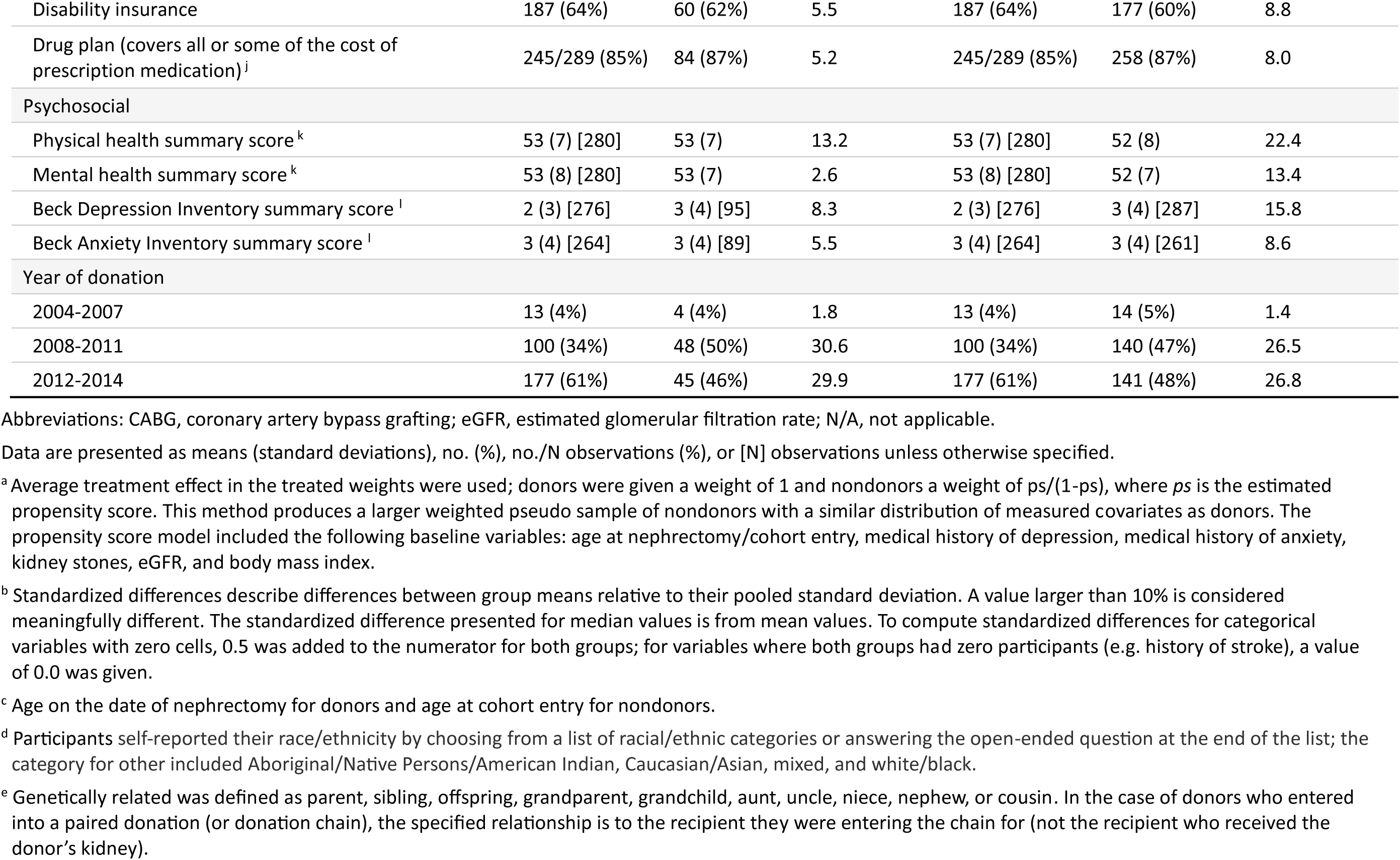
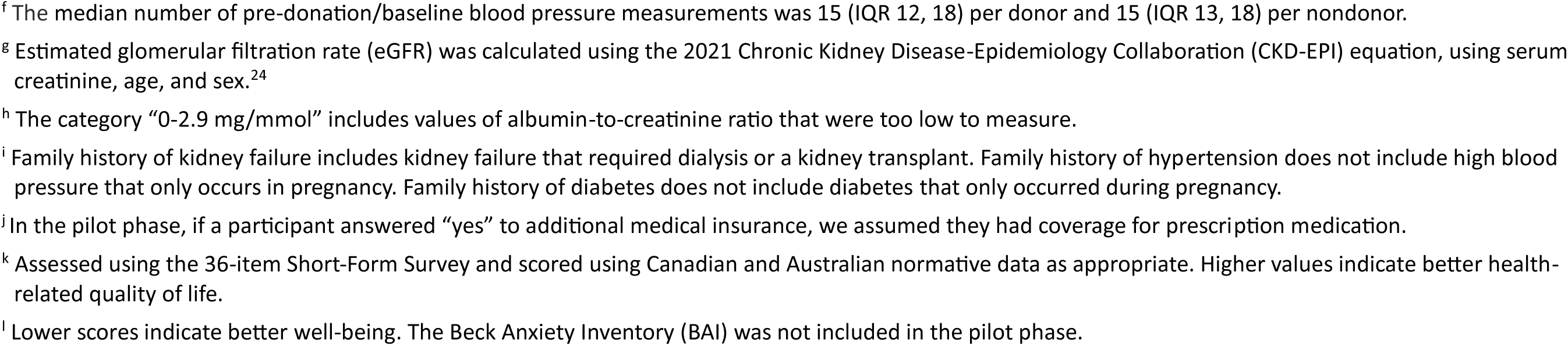
Baseline (pre-donation) characteristics of 290 male living kidney donors and 97 male nondonors before and after weighting.

After statistical weighting, the sample of nondonors increased from 97 to a pseudo sample of 295, with the two groups becoming more balanced on baseline characteristics, including age, BMI, eGFR, a history of kidney stones, a medical history of depression, and a medical history of anxiety.

### Testicular pain in donors vs. nondonors

In the weighted analysis, the risk of testicular pain was significantly greater in donors vs. nondonors: 51/290 (17.6%) vs. 7/295 (2.3%); weighted risk ratio, 7.8 (95% CI, 2.7 to 22.8) (**Table 2**). The two groups did not differ statistically on self-reported hand or eye pain. Effect estimates were similar in the unweighted analysis (**Table 2**).

**Table 2.**
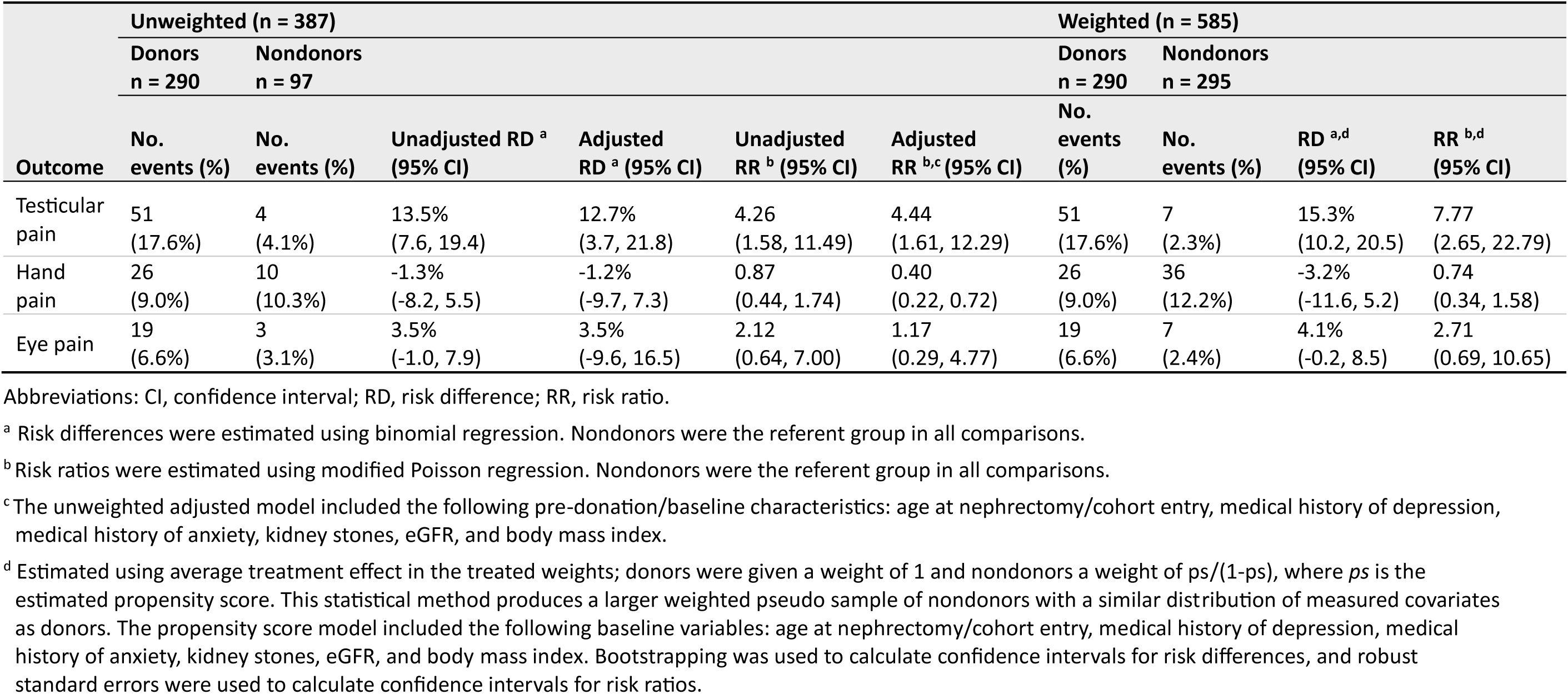
Risk of testicular pain, hand pain, or eye pain in male donors compared with male nondonors.

### Surgical details and testicular pain in donors

Of 290 donors, 246 (84.8%) had laparoscopic surgery, 44 (15.2%) had open surgery, 253 (87.2%) had a left-sided nephrectomy, and 37 (12.8%) had a right-sided nephrectomy. As shown in **Table 3**, left-sided nephrectomies were more likely to be done laparoscopically than right-sided nephrectomies: 88.1% vs. 62.2%, respectively (p<0.001). As shown in **Table 4**, testicular pain was most often unilateral (92.2%) and on the same side as the nephrectomy (ipsilateral) (90.2%).

**Table 3.** Surgical details for 290 male living kidney donors.

| Type of surgery | Nephrectomy side |  | p-value <sup>b</sup> |
| --- | --- | --- | --- |
|  | Left (n=253; 87.2%) | Right (n=37; 12.8%) |  |
| Laparoscopic <sup>a</sup> (n=246; 84.8%) | 223/253 (88.1%) | 23/37 (62.2%) | <0.001 |
| Open (n=44; 15.2%) | 30/253 (11.9%) | 14/37 (37.8%) |  |

<sup>a</sup> Includes hand-assisted laparoscopic surgery. Three of the open surgeries were originally intended to be laparoscopic surgeries
<sup>b</sup> P value computed using a chi-square test.

**Table 4.** Testicular pain by type of surgery and side of nephrectomy in 290 male donors ^a^.

|  | Type of surgery |  |  |  |  |  |
| --- | --- | --- | --- | --- | --- | --- |
|  | Laparoscopic (n=246) |  |  | Open (n=44) |  |  |
|  | Nephrectomy side |  |  | Nephrectomy side |  |  |
| Testicular pain | Left (n=223) | Right (n=23) | Overall (n=246) | Left (n=30) | Right (n=14) | Overall (n=44) |
| Any pain (left or right) (n=51) | 43 (19.3%) | 5 (21.7%) | 48 (19.5%) | 1 (3.3%) | 2 (14.3%) | 3 (6.8%) |
| Pain on left side only (n=41) | 39 (17.5%) | 1 (4.4%) | 40 (16.3%) | 1 (3.3%) | 0 | 1 (2.3%) |
| Pain on right side only (n=6) | 0 | 4 (17.4%) | 4 (1.6%) | 0 | 2 (14.3%) | 2 (4.6%) |
| Pain on both sides (n=4) | 4 (1.8%) | 0 | 4 (1.6%) | 0 | 0 | 0 |

<sup>a</sup> Of 51 donors who reported any testicular pain, unilateral pain was reported by 47 (92.2%), and in 46 (90.2%), it was ipsilateral to the nephrectomy.

When examined by surgery type, testicular pain was more common in donors who had laparoscopic vs. open surgery: 48/246 (19.5%) vs. 3/44 (6.8%); risk difference: 12.7% (95% CI, 3.8% to 21.6%) (**Table 5**). The results were consistent when we only considered ipsilateral testicular pain and when we adjusted for the nephrectomy side, although the confidence intervals were wide.

**Table 5.** Testicular pain after laparoscopic vs. open surgery in 290 male living kidney donors.

| Outcome | Type of surgery |  | Unadjusted risk difference <sup>c</sup> (95% CI) | Adjusted risk difference <sup>c,d</sup> (95% CI) | Unadjusted risk ratio <sup>c</sup> (95% CI) | Adjusted risk ratio <sup>c,d</sup> (95% CI) |
| --- | --- | --- | --- | --- | --- | --- |
|  | Laparoscopic (n=246) <sup>a</sup> | Open (n=44) <sup>b</sup> |  |  |  |  |
| Any testicular pain | 48 (19.5%) | 3 (6.8%) | 12.7% (3.8%, 21.6%) | 15.0% (6.5%, 23.5%) | 2.86 (0.93, 8.78) | 3.08 (1.03, 9.15) |
| Ipsilateral testicular pain | 43 (17.5%) | 3 (6.8%) | 10.7% (1.8%, 19.5%) | 12.8% (4.0%, 21.6%) | 2.56 (0.83, 7.90) | 2.70 (0.91, 8.04) |

Abbreviations: CI, confidence interval.
<sup>a</sup> Laparoscopic includes hand-assisted laparoscopic surgery.
<sup>b</sup> Three of the open surgeries were originally intended to be laparoscopic surgeries.
<sup>c</sup> Risk differences were estimated from a binomial regression model with an identity link, and risk ratios from a modified Poisson regression model.<sup>23</sup>
<sup>d</sup> Adjusted for left- vs. right-sided nephrectomy.

When examined by nephrectomy side, testicular pain was similar in donors who had a left-sided vs. right-sided nephrectomy: 44/253 (17.4%) vs. 7/37 (18.9%), risk difference: −1.5% (95% CI, −15.0% to 11.9%) (**Table 6**). The results were consistent when we only considered ipsilateral testicular pain and when we stratified the analysis by laparoscopic vs. open nephrectomy.

**Table 6.**
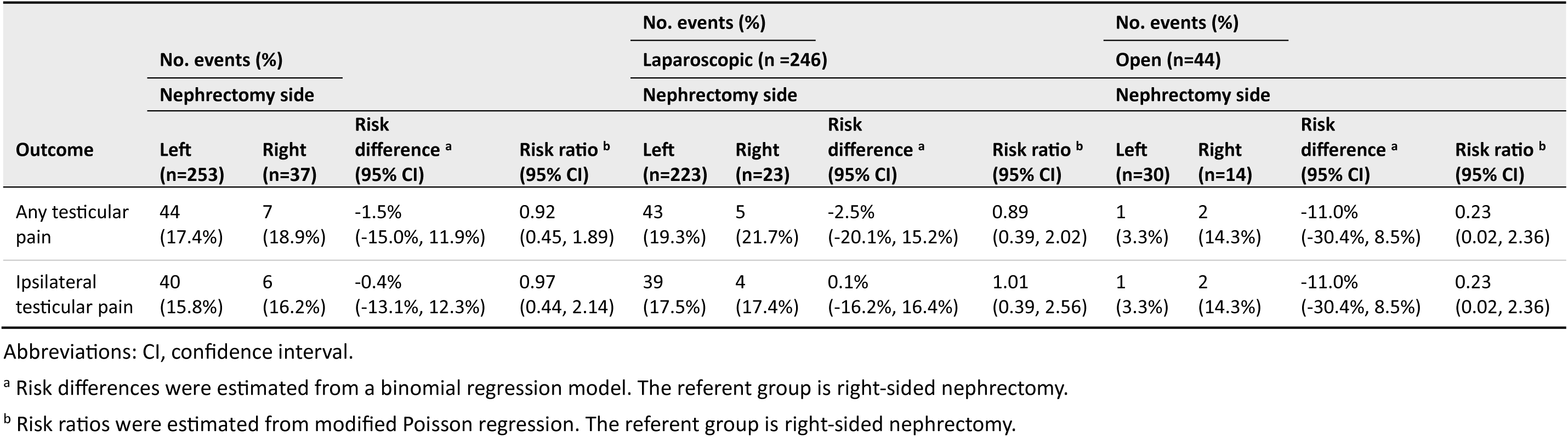
Testicular pain after left-sided vs. right-sided nephrectomy in 290 male living kidney donors, overall and stratified by surgery type.

We reviewed the medical notes for 37 men who had a right-sided nephrectomy. In 13 men, the right gonadal vein was divided; in 22, it remained intact; and in 2, the status was unclear (eTable 3). Out of 35 men (excluding 2 with unclear data), 11 of 21 men who had laparoscopic surgery had the gonadal vein divided (52%), compared to 2 of 14 men who underwent open surgery (14%) (p=0.022). After laparoscopic surgery, right-sided testicular pain occurred in 3 of 11 men who had it divided (27%) compared to 1 of 10 men when it remained intact (10%) (p-value for difference 0.59) (eTable 4). After open surgery, right-sided testicular pain occurred in 1 of 2 men who had it divided (50%) compared to 1 of 12 men when it remained intact (8%) (p-value for difference 0.27).

When examined across surgical centers, testicular pain was reported by donors whose nephrectomies were done in 12 centers located in 6 provinces in Canada and in Australia (eTable 5). Of the 2 centers for which donors did not report testicular pain, 1 center performed 2 nephrectomies, and the other performed 12. Examining the operating room notes, we failed to identify a clear surgical detail to suggest why the center that performed 12 nephrectomies had no men report testicular pain. As shown in eTable 6, the proportion of donors who reported testicular pain was relatively consistent across the study years, and present in more recent years. Pre-donation age, height, weight, and BMI did not differ significantly between donors who reported testicular pain in follow-up compared to those who did not (**Table 7**).

**Table 7.** Pre-donation characteristics and post-donation testicular pain.

| Pre-donation characteristic | Any testicular pain after donation, mean (95% CI) |  |  | Ipsilateral testicular pain, mean (95% CI) |  |  |
| --- | --- | --- | --- | --- | --- | --- |
|  | No (n = 239) | Yes (n = 51) | P-value<sup>a</sup> | No (n = 244) | Yes (n = 46) | P-value<sup>a</sup> |
| Age at donation, years | 49.1 (47.7 to 50.5) | 46.7 (43.6 to 49.7) | 0.16 | 48.9 (47.5, 50.3) | 47.2 (44.0, 50.4) | 0.34 |
| Height, cm | 177.5 (176.6 to 178.3) | 178.1 (176.1 to 180.0) | 0.57 | 177.5 (176.6, 178.3) | 177.9 (175.8, 180.0) | 0.72 |
| Weight, kg | 85.6 (84.1 to 87.1) | 85.7 (82.6 to 88.8) | 0.94 | 85.7 (84.2, 87.1) | 85.3 (81.9, 88.7) | 0.84 |
| Body mass index, kg/m<sup>2</sup> | 27.1 (26.7 to 27.6) | 27.0 (26.2 to 27.9) | 0.80 | 27.2 (26.8, 27.6) | 26.9 (26.0, 27.8) | 0.65 |

Abbreviations: CI, confidence interval.
<sup>a</sup> Computed using a t-test.

Donors provided open-ended responses describing the nature of their testicular pain; symptoms included dull pain, sharp pain, tenderness, aching, and swelling (eTable 7). At least 10 donors indicated they developed a hydrocele in follow-up, and 4 reported having it surgically repaired.

## DISCUSSION

In this cohort study conducted in Canada and Australia, testicular pain was reported by 17.6% of male living kidney donors compared with 2.3% of male nondonors who were statistically weighted to have similar baseline health characteristics as donors. Among donors, testicular pain did not differ between those who had a left-sided vs. right-sided nephrectomy (17.4% vs. 18.9%) but was more likely in those who had laparoscopic vs. open surgery (19.5% vs. 6.8%) even after adjusting for left-vs. right-sided nephrectomy.

The generalizability of our findings is supported by the occurrence of this outcome over a 10-year donation period in multiple provinces in Canada and Australia and by reports of this outcome in 12 other studies from 6 other countries.^9–20^ The proportion of men affected in prior studies varied widely, from 2% to 55%. This variation in estimates is influenced by differing surgical techniques, study designs, and methods of outcome ascertainment. Some studies included a mix of laparoscopic and open nephrectomies,^11,12,14^ while others included only donors who had a left-sided laparoscopic nephrectomy.^10,15,19,20^

In our study, left-sided nephrectomies were more likely to be done laparoscopically and right-sided nephrectomies with an open surgery. In our study and one other,^17^ when only laparoscopic surgeries were considered, the occurrence of testicular pain was similar in donors who had a left-sided vs. right-sided nephrectomy (in our study 19.3% vs. 21.7%, respectively).

In prior studies, the highest rates of testicular pain (>50%) were reported in prospective evaluations,^10,13^ and rates tended to be higher in studies that used direct questioning or surveys rather than chart review.^12,18^ While several studies restricted the outcome definition to ipsilateral pain (testicular pain on the same side as the nephrectomy), contralateral and bilateral testicular pain are also reported after nephrectomy, although less commonly.^17^ In studies that measured onset and duration, testicular pain typically started within 1–4 weeks after nephrectomy and resolved within 1-4 months;^10,17,20^ however, as also observed in our study (etable 7), some donors in prior studies reported pain lasting more than a year.^14,16,18^ On a scale of 1 to 10 (with 10 being the worst possible pain), the pain was rated at an average level of 3–4.^17,20^ In most cases, the pain improved with conservative management (e.g., non-opioid analgesics), but some men were diagnosed with hydroceles and required a hydrocelectomy.^17^ Several donors in our study also indicated they developed a hydrocele in follow-up, and some had it surgically repaired.

In studies that examined whether any pre-donation characteristics predicted post-donation testicular pain, few variables consistently differed between men who experienced pain and those who did not.^16,17^ Older age^11,12^ and taller height^17^ were associated with a greater likelihood of pain in some studies, although these associations were not observed in our study.

Most living donor nephrectomies are now done laparoscopically as this procedure is less invasive and typically offers faster recovery than open surgery. In our study and others, testicular pain was more commonly reported after laparoscopic than open surgery (in our study, the values were 19.5% vs. 6.8%).^12,14^ In other studies, testicular pain after open surgery was reported by 1% to 10% of donors.^11,12,14^

These findings raise questions about why more men experience testicular pain after laparoscopic surgery compared to open surgery and why both procedures cause some pain. In a left-sided nephrectomy, the gonadal vein is always divided, potentially reducing blood flow from the testicle causing venous congestion. Conversely, in a right-sided nephrectomy, the vein may remain intact. In our study, the gonadal vein in right-sided nephrectomies was divided 50% of the time in laparoscopic surgeries compared to 14% with open surgery (eTable 3). The testicular artery is also divided, potentially contributing to ischemic pain, although collateral arterial supply remains from the cremasteric artery and the artery of the vas deferens. To our knowledge, testicular necrosis has not been reported after living donor nephrectomy. However, other factors beyond dividing the gonadal vessels may also contribute to testicular pain. Laparoscopic and open nephrectomies both may disrupt lymphatics, predispose to reflex sympathetic dystrophy from nerve damage to the spermatic plexus and cord, and lead to hydrocele development (fluid accumulation in the tunica vaginalis).^16^ A greater amount of unintentional damage may occur with laparoscopic compared to open surgeries, perhaps related to thermal injuries from the energy devices used in laparoscopic surgery.

Although our study included a nondonor comparison group, it has several important limitations. Participants recalled their symptoms many years after cohort entry, some participants did not complete the survey, and we did not inquire whether participants had a history of scrotal conditions at the time of cohort entry. In addition, we did not assess the timing, severity, or duration of the pain, nor treatments used to treat pain or their response. Our study also lacks information on key surgical details, such as where the gonadal vein was divided.

## Conclusion

Testicular pain may be an underappreciated potential complication of living donor nephrectomy. In this cohort study conducted in Canada and Australia, testicular pain was reported by significantly more male donors during follow-up than male nondonors. Further investigation is needed to characterize this potential complication better and determine if it can be mitigated.

## Supporting information

eTable

## Data Availability

All data produced in the present work are contained in the manuscript.

