## Supplementary material for "Testicular pain after living kidney donation: Results from a multicenter cohort study": eTable

**eTables**


**eTable 1**. STROBE checklist of items that should be included in reports of cohort studies.

**eTable 2**. Study eligibility criteria for nondonors and the screening criteria used to define standard-criteria living kidney donors

**eTable 3.** Gonadal vein clipped and/or divided by surgery type for 37 male donors with a right-sided nephrectomy

**eTable 4.** Testicular pain by type of surgery and whether gonadal vein was clipped and/or divided for 35 donors with a right-sided nephrectomy and non-missing information on whether the gonadal vein was clipped and/or divided

**eTable 5**. Reported testicular pain after living donor nephrectomy across surgery centers, overall and restricted to laparoscopic surgeries

**eTable 6**. Year of donation and reported testicular pain after living donor nephrectomy, overall and restricted to laparoscopic surgeries

**eTable 7**. Donors' descriptions of testicular pain experienced after nephrectomy

**Appendix.** Eye, hand, and testicular pain questionnaire

**eTable 1.** STROBE checklist of items that should be included in reports of cohort studies

| | Item No | Recommendation | Section |
|---|---|---|---|
| **Title and abstract** | 1 | (*a*) Indicate the study's design with a commonly used term in the title or the abstract | Title and abstract |
| | | (*b*) Provide in the abstract an informative and balanced summary of what was done and what was found | Abstract |
| **Introduction** | | | |
| Background/rationale | 2 | Explain the scientific background and rationale for the investigation being reported | Introduction |
| Objectives | 3 | State specific objectives, including any prespecified hypotheses | Introduction |
| **Methods** | | | |
| Study design | 4 | Present key elements of study design early in the paper | Methods |
| Setting | 5 | Describe the setting, locations, and relevant dates, including periods of recruitment, exposure, follow-up, and data collection | Methods |
| Participants | 6 | (*a*) Give the eligibility criteria, and the sources and methods of selection of participants. Describe methods of follow-up | Methods; eTable 2 |
| | | (*b*) For matched studies, give matching criteria and number of exposed and unexposed | n/a |
| Variables | 7 | Clearly define all outcomes, exposures, predictors, potential confounders, and effect modifiers. Give diagnostic criteria, if applicable | Methods |
| Data sources/ measurement | 8* | For each variable of interest, give sources of data and details of methods of assessment (measurement). Describe comparability of assessment methods if there is more than one group | Methods Protocol |
| Bias | 9 | Describe any efforts to address potential sources of bias | Methods Protocol[a] |
| Study size | 10 | Explain how the study size was arrived at | Protocol[a] |
| Quantitative variables | 11 | Explain how quantitative variables were handled in the analyses. If applicable, describe which groupings were chosen and why | Methods |
| Statistical methods | 12 | (*a*) Describe all statistical methods, including those used to control for confounding | Methods |
| | | (*b*) Describe any methods used to examine subgroups and interactions | Methods |
| | | (*c*) Explain how missing data were addressed | Results, Figure 1 |
| | | (*d*) If applicable, explain how loss to follow-up was addressed | Protocol[a] |
| | | (*e*) Describe any sensitivity analyses | Methods |
| **Results** | | | |
| Participants | 13* | (a) Report numbers of individuals at each stage of study—eg numbers potentially eligible, examined for eligibility, confirmed eligible, included in the study, completing follow-up, and analysed | Results Figure 1 |
| | | (b) Give reasons for non-participation at each stage | Results Figure 1 |
| | | (c) Consider use of a flow diagram | Figure 1 |
| Descriptive data | 14* | (a) Give characteristics of study participants (eg demographic, clinical, social) and information on exposures and potential confounders | Results Table 1 |
| | | (b) Indicate number of participants with missing data for each variable of interest | Results Figure 1 |
| | | (c) Summarise follow-up time (eg, average and total amount) | Results |

| | Item No | Recommendation | Section |
|---|---|---|---|
| Outcome data | 15* | Report numbers of outcome events or summary measures over time | Results |
| Main results | 16 | (*a*) Give unadjusted estimates and, if applicable, confounder-adjusted estimates and their precision (eg, 95% confidence interval). Make clear which confounders were adjusted for and why they were included | Results |
| | | (*b*) Report category boundaries when continuous variables were categorized | N/A |
| | | (*c*) If relevant, consider translating estimates of relative risk into absolute risk for a meaningful time period | Results |
| Other analyses | 17 | Report other analyses done—eg analyses of subgroups and interactions, and sensitivity analyses | Results Supplement |
| **Discussion** | | | |
| Key results | 18 | Summarise key results with reference to study objectives | Discussion |
| Limitations | 19 | Discuss limitations of the study, taking into account sources of potential bias or imprecision. Discuss both direction and magnitude of any potential bias | Discussion |
| Interpretation | 20 | Give a cautious overall interpretation of results considering objectives, limitations, multiplicity of analyses, results from similar studies, and other relevant evidence | Discussion |
| Generalisability | 21 | Discuss the generalisability (external validity) of the study results | Discussion |
| **Other information** | | | |
| Funding | 22 | Give the source of funding and the role of the funders for the present study and, if applicable, for the original study on which the present article is based | Article Information |

*Give information separately for exposed and unexposed groups.

**Note:** An Explanation and Elaboration article discusses each checklist item and gives methodological background and published examples of transparent reporting. The STROBE checklist is best used in conjunction with this article (freely available on the Web sites of PLoS Medicine at http://www.plosmedicine.org/, Annals of Internal Medicine at http://www.annals.org/, and Epidemiology at http://www.epidem.com/). Information on the STROBE Initiative is available at http://www.strobe-statement.org.

[a] Garg AX, Arnold JB, Cuerden M, et al. The Living Kidney Donor Safety Study: Protocol of a Prospective Cohort Study. *Can J Kidney Health Dis*. 2022;9. doi:10.1177/20543581221129442

**eTable 2.** Study eligibility criteria for nondonors and the screening criteria used to define standard-criteria living kidney donors*[a]

| **Inclusion criteria**[a] | |
|---|---|
| Age | Age between 18 to 70 years. |
| Blood pressure | Average systolic/diastolic blood pressure <140/90 mm Hg based on an average of at least three blood pressure measurements taken during the recruitment interview. If the average of these blood pressure measurements was elevated, the participant was still eligible for participation if the average of an additional 12 home blood pressure readings was <140/90 mm Hg. All participants need to successfully record at least 12 home blood pressure readings using the self-monitoring device to be eligible. |
| Kidney function | Serum creatinine <115 µmol/L in men or <90 µmol/L in women[2] or a Cockcroft-Gault estimated glomerular filtration rate >80 mL/min. |
| No urine protein | Negative urine dipstick for protein, or if trace or 0.3 g/L, a random urine albumin-to-creatinine ratio <8 mg/mmol. |
| No hematuria | Negative urine dipstick for hematuria. Those with non-persistent hematuria are eligible to participate; those with initial evidence of dipstick hematuria may have a second assessment, and for women this should not occur during the time of menses. Individuals with hematuria that resolves after treatment of a urinary tract infection are eligible for study participation.[3] |
| Non-obese body mass index | Body mass index <35 kg/m$^2$. |
| Language | Ability to speak and read English or French. |
| **Exclusion criteria**[a] | |
| Anti-hypertensive medication | Taking anti-hypertensive medication on a daily basis for any reason. |
| Kidney stones | Symptoms or evidence of kidney stones in the past 3 years. |
| History of kidney failure | Recipient of a kidney transplant; ever received dialysis. |
| Elevated plasma glucose or history of gestational diabetes | Plasma glucose ≥7.0 mmol/L after a 6-hour fast (if available); 2-hour oral glucose ≥11.1 mmol/L (if available). History of gestational diabetes. |
| Comorbidities | History of kidney disease; cancer, other than cured non-melanoma skin cancer; diabetes; cardiovascular disease; or pulmonary disease. |
| Contraindications to living kidney donation, general anesthesia, or surgery | Has a medical condition that would prevent them from becoming a living kidney donor or a known contraindication to general anesthesia or surgery. |
| Pregnancy | Currently pregnant, pregnant in the last month, or planning to become pregnant in the coming year. |
| Other study participation | Participating in a clinical trial or another study that could influence the outcomes of this study. |

*First published by SAGE under a Creative Commons Attribution-Non-Commercial 4.0 License (Garg et al., The Living Kidney Donor Safety Study: Protocol of a Prospective Cohort Study. *Can J Kid Health Dis*. 2022. doi.org/10.1177/20543581221129442).

[a] Donor candidates were automatically eligible to participate in the study if they were approved for donation by their local transplant center, were able to speak and read English and/or French, and were not participating in a clinical study that would affect the outcome of this study. Prospective donors who were approved for donation but who did not meet these screening criteria were deemed expanded-criteria donors. To be eligible to participate in this study, nondonors had to meet the same screening criteria as standard-criteria donors.

**eTable 3.** Gonadal vein clipped and/or divided by surgery type for 37 male donors with a right-sided nephrectomy

| | Type of surgery [a] | | |
|---|---|---|---|
| **Gonadal vein clipped and/or divided** | **Laparoscopic (n=23)** | **Open (n=14)** | **Overall (n=37)** |
| Yes | 11/21 (52%) | 2 (14%) | 13/35 (37%) |
| No | 10/21 (48%) | 12 (86%) | 22/35 (63%) |
| Unsure | 2 (9%) | 0 | 2 (5%) |

[a] The chi-square test p-value for testing whether donors undergoing laparoscopic right-sided nephrectomy are more likely to have the gonadal vein clipped and/or divided compared to open surgery is 0.022.

**eTable 4.** Testicular pain by type of surgery and whether gonadal vein was clipped and/or divided for 35 donors with a right-sided nephrectomy and non-missing information on whether the gonadal vein was clipped and/or divided

| | | | | | Laparoscopic (n=21) | | | | Open (n=14) | | | |
|---|---|---|---|---|---|---|---|---|---|---|---|---|
| | No. events (%) | | | | No. events (%) | | | | No. events (%) | | | |
| | Gonadal vein | | | | Gonadal vein | | | | Gonadal vein | | | |
| Testicular pain | Clipped and/or divided (n=13) | Preserved (n=22) | Risk difference [a] (95% CI) | Risk ratio [b] (95% CI) | Clipped and/or divided (n=11) | Preserved (n=10) | Risk difference [a] (95% CI) | Risk ratio [b] (95% CI) | Clipped and/or divided (n=2) | Preserved (n=12) | Risk difference [a] (95% CI) | Risk ratio [b] (95% CI) |
| Any pain | 4 (31%) | 3 (14%) | 17.1% (-11.7%, 46.0%) | 2.26 (0.60, 8.54) | 3 (27%) | 2 (20%) | 7.3% (-28.9%, 43.4%) | 1.36 (0.28, 6.56) | 1 (50%) | 1 (8%) | 41.7% (-29.4%, 112.7%) | 6.00 (0.58, 61.84) |
| Ipsilateral | 4 (31%) | 2 (9%) | 21.7% (-6.1%, 49.5%) | 3.38 (0.72, 16.0) | 3 (27%) | 1 (10%) | 17.3% (-15.0%, 49.5%) | 2.73 (0.34, 22.16) | 1 (50%) | 1 (8%) | 41.7% (-29.4%, 112.7%) | 6.00 (0.58, 61.84) |

Abbreviations: CI, confidence interval.

[a] Risk differences were estimated from a binomial regression model. The referent group is right-sided nephrectomy.

[b] Risk ratios were estimated from modified Poisson regression. The referent group is right-sided nephrectomy.

**eTable 5.** Reported testicular pain after living donor nephrectomy across surgery centers, overall and restricted to laparoscopic surgeries

| Surgery Center | Any testicular pain | | Ipsilateral testicular pain | |
|---|---|---|---|---|
| | All surgeries (n = 290) | Laparoscopic only [a] (n = 246) | All surgeries (n = 290) | Laparoscopic only [a] (n = 246) |
| **Canada** | | | | |
| *Alberta* | | | | |
| Calgary – Foothills Medical Centre | 2/11 (18%) | 2/9 (22%) | 2/11 (18%) | 2/9 (22%) |
| Edmonton – University of Alberta Hospital | 4/29 (14%) | 1/12 (8%) | 4/29 (14%) | 1/12 (8%) |
| *British Columbia* | | | | |
| Vancouver – St. Paul's Hospital | 10/49 (20%) | 10/49 (20%) | 9/49 (18%) | 9/49 (18%) |
| Vancouver – Vancouver General Hospital | 6/25 (24%) | 6/25 (24%) | 5/25 (20%) | 5/25 (20%) |
| *Nova Scotia* | | | | |
| Halifax – Queen Elizabeth II Health Sciences Centre | 0/12 (0%) | 0/12 (0%) | 0/12 (0%) | 0/12 (0%) |
| *Manitoba* | | | | |
| Winnipeg – Health Sciences Centre | 3/10 (30%) | 3/6 (50%) | 3/10 (30%) | 3/6 (50%) |
| *Ontario* | | | | |
| Hamilton – St. Joseph's Healthcare | 2/5 (40%) | 2/5 (40%) | 1/5 (20%) | 1/5 (20%) |
| London – University Hospital | 6/38 (16%) | 6/37 (16%) | 6/38 (16%) | 6/37 (16%) |
| Ottawa – The Ottawa Hospital | 5/32 (16%) | 5/31 (16%) | 4/32 (13%) | 4/31 (13%) |
| Toronto – St. Michael's Hospital | 3/17 (18%) | 3/17 (18%) | 3/17 (18%) | 3/17 (18%) |
| Toronto – University Health Network | 4/33 (12%) | 4/16 (25%) | 3/33 (9%) | 3/16 (19%) |
| *Quebec* | | | | |
| Montreal – Notre Dame Hospital | 0/2 (0%) | 0/2 (0%) | 0/2 (0%) | 0/2 (0%) |
| Montreal – Royal Victoria Hospital | 3/9 (33%) | 3/9 (33%) | 3/9 (33%) | 3/9 (33%) |
| **Australia** [b] | 3/18 (17%) | 3/16 (19%) | 3/18 (17%) | 3/16 (19%) |

[a] Laparoscopic includes hand-assisted laparoscopic, robotic-assisted laparoscopic, and straight laparoscopic. Surgeries that were planned laparoscopic and converted to open were categorized as open (n=3).

[b] Includes 3 surgical centers.

**eTable 6.** Year of donation and reported testicular pain after living donor nephrectomy, overall and restricted to laparoscopic surgeries

| Year of donation | Any testicular pain | | Ipsilateral testicular pain | |
|---|---|---|---|---|
| | **All surgeries (n = 290)** | **Laparoscopic only [a] (n = 246)** | **All surgeries (n = 290)** | **Laparoscopic only [a] (n = 246)** |
| 2004 – 2009 [b] | 2/20 (10%) | 2/19 (11%) | 1/20 (5%) | 1/19 (5%) |
| 2010 | 6/30 (20%) | 5/23 (22%) | 5/30 (17%) | 4/23 (17%) |
| 2011 | 10/63 (16%) | 9/51 (18%) | 10/63 (16%) | 9/51 (18%) |
| 2012 | 10/69 (14%) | 9/59 (15%) | 10/69 (14%) | 9/59 (15%) |
| 2013 | 14/70 (20%) | 14/59 (24%) | 13/70 (19%) | 13/59 (22%) |
| 2014 | 9/38 (24%) | 9/35 (26%) | 7/38 (18%) | 7/35 (20%) |

[a] Laparoscopic includes hand-assisted laparoscopic, robotic-assisted laparoscopic, and straight laparoscopic. Surgeries that were planned laparoscopic and converted to open were categorized as open (n=3).

[b] The study's pilot phase occurred between 2004–2008, and most donors in the pilot phase were not included in the main study. The proportion of donors who reported testicular pain was not statistically different between those who donated from 2004–2009 vs. from 2010–2014 (chi-square test; p=0.36); similarly, the proportion who reported ipsilateral testicular pain was not statistically different between these time periods (chi-square test; p=0.17).

**eTable 7**. Donors' descriptions of testicular pain experienced after nephrectomy

| Pain/tenderness/numbness/pressure | Hydroceles | Swelling |
|---|---|---|
| • Medium pressure in testicle, periodically.<br>• Occasionally I have left testicular pain.<br>• Sharp pain in groin migrates to left abdomen - intermittent pain when lying on left.<br>• Sensitivity to pressure.<br>• Pressure in left testicle; consulted doctor with ultrasound and found a scrotal pearl.<br>• Left testicle sore since kidney donation.<br>• Tender to touch. Tenderness noticeable when crossing legs, etc.<br>• Tenderness. Family physician diagnosed as normal and transient.<br>• Soreness for 3-4 days several months apart. Symptoms resolved with time. No recurrence.<br>• Mild discomfort in the left testicle and above the left pubic area.<br>• The pain is mild ... a dull ache ... I feel it only when touched there, or when pressure is applied there.<br>• Dull ache.<br>• Periodic aching.<br>• Pain since recovery. Sore for 6 months had an investigation done.<br>• Numbness - feels almost like it is asleep (ever since surgery).<br>• An achiness in left testicle - could feel the ache a little in my groin/pelvic region.<br>• Sharp pain in left testicle then goes away fast.<br>• Sharp/throbbing.<br>• A dull ache most days. Very sensitive to touch.<br>• Dull ache it doesn't last long and is infrequent.<br>• Post surgery - aching. 3 weeks. The surgeon said it's a result of the surgery and would go away. It did.<br>• It used to constantly hurt, but now it only seems sensitive to any kind of contact.<br>• Constant pain in my left testicle.<br>• The pain is a sharp stabbing pain at different times.<br>• Massive swelling and pain post surgery for a few weeks. Not once was warned this could happen during donor assessment. Still sensitive to this day and a sudden pressure can cause pain esp in left (ie cat walking on lap). | • Diagnosed with hydrocele - left testicle. More like discomfort - not pain.<br>• Since my left kidney was removed for donation I have developed a left testicular hydrocele. I had a testicular ultrasound done last year and it was reviewed by the nephrologist who did my surgery and felt it could be observed for now.<br>• Hydrocele surgery.<br>• Had hydrocele repaired, which solved the pain.<br>• Hydrocele removed after surgery.<br>• Hydrocele formed 3 months after surgery - CT scan shows about 25 ml of fluid buildup.<br>• Doctors identified it as a hydrocele.<br>• Throbbing. It's a hydrocele. Caused by kidney donation.<br>• Hydrocele in right testicle, surgery was required.<br>• Extremely sensitive for 1 year post surgery - 8 years later - 5 lb hydrocele cyst - reduced in size prior to surgery - so did not have the surgery. | • Swollen testicles; saw resident at unit.<br>• No pain - doubled in size.<br>• Swollen, tender to touch.<br>• Swollen scrotum after donation; 3x previous size; seen by urologist.<br>• Sensitive to touch, similar to a bruise. Left testicle is swollen.<br>• I noticed after surgery that my left testicle got very large. It is much more sensitive but not painful. But it does worry me.<br>• Left testicle has enlarged.<br>• Dilation of the testicle veins. |

### Appendix

LKD STUDY 3 - ☐☐☐ - ☐☐☐ - 1 Participant Initials: ☐☐☐ Assessment #: ☐☐ Form #: 4 9

Centre ID Participant ID F M L

**PAIN FORM**

1.) Since ____________________, have you experienced any **NEW** pain in the following areas?

**PLEASE CHECK ALL THAT APPLY:**

☐ Eyes

If yes, which side did the pain occur?

- ☐ Left
- ☐ Right
- ☐ Both

☐ Hands

If yes, which side did the pain occur?

- ☐ Left
- ☐ Right
- ☐ Both

☐ Testicles

If yes, which side did the pain occur?

- ☐ Left
- ☐ Right
- ☐ Both

2.) Please, describe the pain you feel or felt?

______________________________________________

______________________________________________

______________________________________________

______________________________________________

______________________________________________

Person completing form (please print): ______________________ ________

last name first initial

Version: 30 January 2014 Page 1 of 1
